# Immunogenicity and reactogenicity after booster dose with AZD1222 via intradermal route among adult who had received CoronaVac

**DOI:** 10.1101/2021.12.12.21267695

**Authors:** Rapisa Nantanee, Puneyavee Aikphaibul, Peera Jaru-Ampornpan, Pimpayao Sodsai, Orawan Himananto, Tuangtip Theerawit, Jiratchaya Sophonphan, Punyot Tovichayathamrong, Kasama Manothummetha, Tysdi Laohasereekul, Narin Hiransuthikul, Nattiya Hirankarn, Thanyawee Puthanakit, on behalf of the Study Team

**Affiliations:** Center of Excellence in Pediatric Infectious Diseases and Vaccines, Chulalongkorn University, 1873, Rama IV Rd, Pathumwan, Bangkok, 10330, Thailand; Pediatric Allergy and Clinical Immunology Research Unit, Division of Allergy and Immunology, Department of Pediatrics, Faculty of Medicine, Chulalongkorn University, King Chulalongkorn Memorial Hospital, The Thai Red Cross Society, 1873, Rama IV Rd, Pathumwan, Bangkok, 10330, Thailand; Division of Pediatric Dermatology, Department of Pediatrics, Faculty of Medicine, Chulalongkorn University, King Chulalongkorn Memorial Hospital, The Thai Red Cross Society, 1873, Rama IV Rd, Pathumwan, Bangkok, 10330, Thailand; Virology and Cell Technology Research Team, National Center for Genetic Engineering and Biotechnology (BIOTEC); Center of Excellence in Immunology and Immune-mediated Diseases, Department of Microbiology, Faculty of Medicine, Chulalongkorn University, 1873, Rama IV Rd, Pathumwan, Bangkok, 10330, Thailand; Monoclonal Antibody Production and Application Research Team, National Center for Genetic Engineering and Biotechnology (BIOTEC); The HIV Netherlands Australia Thailand Research Collaboration (HIV-NAT), The Thai Red Cross AIDS Research Centre, Bangkok, Thailand; Department of Microbiology, Faculty of Medicine, Chulalongkorn University, 1873, Rama IV Rd, Pathumwan, Bangkok, 10330, Thailand; Department of Preventive and Social Medicine, Faculty of Medicine, Chulalongkorn University, 1873, Rama IV Rd, Pathumwan, Bangkok, 10330, Thailand

**Author notes:** Corresponding author: Thanyawee Puthanakit, MD Center of Excellence in Pediatric Infectious Diseases and Vaccines, Department of Pediatrics, Faculty of Medicine, Chulalongkorn University, 1873, Rama IV Rd., Pathumwan, Bangkok, 10330, Thailand. Additional study team members are listed (on behalf of the Study Team).

**Keywords:** SARS-CoV2 vaccine, booster dose, AZD1222, Neutralizing antibody titer, anti-SARS CoV2 IgG, CoronaVac vaccine, ChAdOx1 nCoV-19 vaccine, intradermal

## Abstract

**Background:** Currently, booster dose is needed after 2 doses of inactivated COVID-19 vaccine. With limited resource and shortage of COVID-19 vaccine, intradermal(ID) administration might be a potential dose-sparing strategy.

**Objective:** To determine antibody response and reactogenicity of ID ChAdOx1 nCoV-19 vaccine(AZD1222,Oxford/AstraZeneca) as a booster dose after completion of 2-dose CoronaVac(SV) in healthy adult.

**Methods:** This is a prospective cohort study of adult aged 18-59 years who received 2-dose SV at 14-35 days apart for more than 2 months. Participants received ID AZD1222 at fractional low dose(1×10^10^ viral particles,0.1ml). Antibody responses were evaluated by surrogate virus neutralization test(sVNT) against wild type and delta variant and anti-spike-receptor-binding-domain immunoglobulin G(anti-S-RBD IgG) at prior, day14 or 28, and day90 post booster. Solicited reactogenicity was collected during 7 days post-booster. Primary endpoint was the differences of sVNT against delta strain ≥80%inhibition at day14 and 90 compared with the parallel cohort study of 0.5-ml intramuscular(IM) route.

**Results:** From August2021, 100 adults with median(IQR) age of 46(41-52) years participated. At baseline, geometric means(GMs) of sVNT against delta strain prior to booster were 22.4%inhibition(95%CI 18.7-26.9) and of anti-S-RBD IgG were 109.3(95.4-125.1)BAU/ml. GMs of sVNT against delta strain were 92.9%inhibition(95%CI 87.7-98.3) at day14 and 73.1%inhibition(66.7-80.2) at day90 post ID booster. The differences of proportion of participants with sVNT to delta strain≥80%inhibition in ID recipients versus IM were +4.2%(95%CI-2.0to10.5) at day14, and -37.3%(−54.2to−20.3) at day90. Anti-S-RBD IgG GMs were 2037.1(95%CI1770.9-2343.2) at day14 and 744.6(650.1-852.9) BAU/ml at day90, respectively. Geometric mean ratios(GMRs) of anti-S-RBD IgG were 0.99(0.83-1.20) at day14, and 0.82(0.66-1.02) at day90. Only 18% reported feverish, compared with 37% of IM(p=0.003). Only 18% reported feverish, compared with 37% of IM(p=0.003). Common reactogenicity was erythema(55%) at injection site while 7% reported blister.

**Conclusion:** Low-dose ID AZD1222 booster enhanced lower neutralizing antibodies at 3 months compared with IM route. Less systemic reactogenicity occurred, but higher local reactogenicity.

**Highlights:**

- Intradermal AZD1222 booster vaccine gave comparable short-term immunogenicity but lower 90-day immunogenicity with conventional intramuscular vaccine.
- Lower systemic but higher local reactogenicity was found in intradermal AZD1222 booster vaccine.
- Blister and pruritus could be seen after intradermal AZD1222 booster vaccine.

## 1. Introduction

Global report of Coronavirus disease (COVID-19) infection was over 250 million cases with more than 5 million deaths [1], despite over 7 billion doses of vaccination have been administered. In Thailand, as of November 2021, more than 2 million reported COVID-19 infection with over 20,000 deaths. Inactivated COVID-19 vaccine, CoronaVac (Sinovac Life Sciences, Beijing, China), was used for mass vaccination in several countries e.g., Thailand, China, Brazil, and Chile. Effectiveness of CoronaVac for prevention of COVID-19 was 65.9% from study in Chile [2] and 36.8% from study in Brazil [3]. With the rising of delta variant (B.1.617.2) of SAR-CoV-2 globally, the neutralizing activity induced by CoronaVac declined [4]. Hence, heterologous prime boost vaccination may provide better immunogenicity. With AZD1222 followed by BNT162b2 heterologous prime boost vaccination, this vaccination strategy provided highest T cell reactivity compared with homologous vaccination [5, 6].

Standard administration of currently available COVID-19 vaccine is via intramuscular injection. Routes for vaccine administration could be intramuscular (IM), intradermal (ID) in which efficacy is related to the immunogenicity [7]. ID administration offers potential dose-sparing benefit compared with intramuscular administration, rabies vaccination as example. ID vaccination is a technique in which the vaccine is administered into dermis which is rich in antigen presenting cells such as dermal dendritic cells [8]. Because of the abundance of antigen presenting cells in skin, ID administration required less antigenic dose (usually 20%-30% of standard dose) to induce immune responses than standard IM vaccination. Many studies showed effective immune response by ID vaccination including influenza, rabies, hepatitis B, Bacille Calmette-Guerin (BCG), and polio vaccination [7, 9–12]. For influenza vaccine, a systematic review and meta-analysis showed comparable seroprotection rates for 9-µg ID with 15-µg IM injection with higher local adverse event particularly erythema and swelling [9]. For rabies vaccine, ID schedules offer advantages through savings in costs, doses and time as recommended by WHO and also approved use on label of vaccine [12].

Fractionated-dose ID COVID-19 vaccine is the potential for rapid achievement of herd immunity based on other vaccines reported [7]. Study of one-tenth dose of mRNA-1273 ID vaccination showed comparable anti-spike IgG and anti-RBD IgG responses to conventional IM vaccination at 2 weeks post primary vaccination series [13]. However, one-fifth dose of BNT162b2 ID booster in healthy Thai adult post 2-dose CoronaVac failed to boost T cell response at 14 days, despite robust neutralizing antibodies response [14]. A case report of ID AZD1222 after 2 doses of CoronaVac showed increase of antibodies and T cell responses against spike protein which neutralizing antibody increased to almost 100% with minimal local reaction at 2-3 weeks after booster [15].

This study aims to evaluate immunogenicity and reactogenicity of ID AZD1222 booster dose in adults who had received 2 doses of CoronaVac.

## 2. Methods

### 2.1 Study design and participants

This study was conducted at Chulalongkorn University Health Center, Faculty of Medicine, Chulalongkorn University, Bangkok Thailand. This is a prospective cohort study. The participants who aged 18 – 59 years old and received two doses of CoronaVac for at least 60 days, 14 – 35 days apart were included in this study. The exclusion criteria were receiving any immunosuppressants or blood products within 3 months before the enrollment or receiving any vaccines within 2 weeks. All participants gave written informed consent prior to study enrollment.

This study was registered in Thai Clinical Trials Registry (thaiclinicaltrials.org, TCTR 20210817003). Immunogenicity parameters were compared with the parallel randomized controlled trial on healthy adult with standard dose and low dose IM administration of AZD1222 booster after completing 2 doses of CoronaVac (TCTR20210722003), conducted at same settings and lab. We compared the results of this study with conventional standard dose (0.5 ml) IM group. Institutional review board of Faculty of Medicine, Chulalongkorn University approved this study (IRB no. 663/64) and parallel IM AZD1222 booster study (IRB no. 600/64).

### 2.2 Study procedures

One hundred participants were recruited in this study. At baseline, the history of SARS-CoV-2 vaccination and exposure to confirmed case within 3 months were taken. Blood sample was collected prior to giving a booster dose. The participants received ID AZD1222 lot number A1009, manufactured by Siam Bioscience Co., Ltd., 0.1 ml (1 x 10^10^ viral particles). The vaccination was performed by trained physician/nurse (RN and TT). The solicited local and systemic reactogenicity during 7 days after vaccination was recorded in the diary. The solicited reactogenicity included fever, pain, swelling, erythema, headache, malaise, myalgia, arthralgia, vomiting, and diarrhea. Scheduled visits were day 14 for 50 participants, day 28 for 50 participants. At day 90, 40 participants were scheduled, to collect reactogenicity data and perform blood collection.

The cell-mediated immunity (CMI) sub study was performed among 20 participants, at baseline, day 28, and day 90, with enzyme-linked immunospot (ELISpot) assay to evaluate T and B cell responses.

### 2.3 Immunogenicity outcomes

All participants’ samples were tested for spike receptor binding domain (S-RBD) IgG, and functional neutralizing antibody (NAb) against SARS-CoV-2 wild type and delta variants by surrogate virus neutralization test (sVNT). All of the immunogenicity results in ID group were compared with IM participants at equivalent time points.

#### Quantitative spike receptor binding domain IgG (anti-S-RBD IgG) ELISA

The ELISA protocol was adapted from Amanat *et al*. (2020) [16]. Briefly, diluted serum samples were incubated in 96-well plates coated with purified recombinant Myc-His-tagged S-RBD, residues 319-541 from SARS-CoV-2 (Wuhan-Hu-1). Then, ELISA was performed. Anti-S-RBD IgG level was reported in binding-antibody units (BAU/mL) following conversion of OD450 values with the standard curve using known units of WHO international standard (NIBSC 20/136). We used anti-S-RBD IgG level at 506 BAU/ml, which is correlated with 80% vaccine efficacy reported by the Oxford COVID vaccine trial group [17], as a cut off.

#### Surrogate virus neutralization test (sVNT)

A surrogate virus neutralization test was set up as previously described in Tan *et al*. (2020) [18]. Recombinant SRBD from the wild-type (Wuhan-Hu-1) and delta (B.1.617.2) strains were used. Serum samples - SRBD mixture were incubated in 96-well plates coated with 0.1 µg/well recombinant human ACE2 ectodomain (GenScript). Then, ELISA was performed. The negative sample was pre-2019 human serum. The % inhibition was calculated as follows:

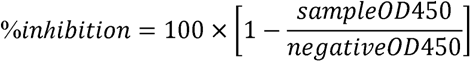

#### Enzyme-linked immunospot (ELISpot) assay to evaluate T and B cell responses

For T cell, ELISpot assay using a Human IFN-γ ELISpotPro^TM^ kit (Mabtech, Stockholm, Sweden) was used for SARS-CoV-2-specific T cell responses in fresh peripheral blood mononuclear cells (PBMCs). Briefly, 2.5 × 10^5^ PBMCs were stimulated in AIM-V medium with overlapping peptide pool from 100 peptides of SARS-CoV-2 Spike (S) defined peptides and 101 peptides from the nucleoprotein (N), membrane protein (M), open reading frame proteins (O) (Mabtech, Stockholm, Sweden) at a final concentration of 2 µg/ml for 20 hours. Negative control and positive control, anti-CD3, were also included. The spots were counted using ImmunoSpot analyzer. Spot counts for negative control wells were subtracted from the test wells to generate normalized readings, these are presented as spot forming unit (SFU) per million PBMCs.

For B cells, Human IgG SARS-CoV-2 RBD ELISpot PLUS (ALP) kit (Mabtech, Stockholm, Sweden) was used for SARS-CoV-2-specific B cell responses. Briefly, the memory B cells were differentiated into antibody secreting cells by pre-stimulating the fresh PBMCs with R848 and IL-2 for 72 hours. Unstimulated well was also used as negative control. Stimulated and unstimulated PBMCs (5 × 10^5^ cells per well) were added into ELISpot plate and incubated for 18 hours. An RBD-WASP antigen was added into RBD-specific IgG detected well while MT78/145-biotinylated antibodies were added into total IgG detected well, positive control. Anti-WASP-ALP was added into RBD-specific IgG detected well and negative control well while streptavidin-ALP was added into total IgG detected well. Spot counting was performed in the same method as T cells.

### 2.4 Reactogenicity

Solicited reactogenicity was recorded by participants using diary. All symptoms were graded in 3 grades [19]: grade 0 for no symptom; grade 1 for mild symptom, which was not interfere with activities or vomiting 1 – 2 times/day or diarrhea 2 – 3 times/day; grade 2 for moderate symptom, which interfered with activities or need to take medication, or vomiting more than 2 times/day or diarrhea 4 – 5 times/day; grade 3 for severe symptom, which incapacitated or need hospitalization or diarrhea 6 or more times/day. Fever was graded as grade 1 (38.0 – 38.4°C), grade 2 (38.5 – 38.9°C), grade 3 (39 – 40°C), and grade 4 (more than 40°C). Unsolicited adverse events were also recorded at all visits by study team.

### 2.5 Statistical analysis

Demographic and clinical characteristics were described for the subjects. Continuous variables were expressed as median (interquartile range: IQR) and number with percentage for categorical variables. Differences in continuous and categorical variables between two groups were assessed using a Wilcoxon rank sum test, Chi-square test, or fisher exact test, respectively. The sVNT results, to either wild type or delta strain, of more than 80% were used to classify the achievement of 80% protection against symptomatic infection. The anti-S-RBD IgG of more than 506 BAU/ml were used in this study as a cut off for protective antibody level, which previously reported to be associated with 80% vaccine efficacy against primary symptomatic COVID-19 [20].

We presented primary comparisons between ID and IM group in terms of the differences of proportion of participants achieving sVNT ≥80%inhibition. Non-inferiority was concluded if the lower bound of the 95% CI did not exceed -10%. Geometric means (GMs) and geometric mean ratios (GMRs) with 95% confidence interval of anti-S-RBD IgG and sVNT, at day 0, 14, 28, and 90 after booster vaccination, were calculated. Non-inferiority was concluded if the lower bound of the 95% CI did not exceed 0.67 for the GMR of anti-S-RBD. All P-values reported are two-sided. Statistical significance was defined as P<0.05. Stata version 15.1 (Stata Corp., College Station, Texas), was used for analysis.

## 3. Results

### 3.1 Baseline characteristic

The participants were enrolled during August 2021. The demographic data was shown in Table 1. Median age was 46 (IQR 41-52) years, 55% were male. Underlying disease was described, as shown in Table 1. Duration between 2 doses of CoronaVac was 21 days with median 71 (IQR 65-76) days prior to ID booster administration. The sVNT against delta strain GM was 44.5% inhibition and against wild type was 22.4% inhibition. The number of female were higher in IM cohort, while the interval between completion of 2-dose CoronaVac and AZD1222 booster was longer in ID than IM cohort.

**Table 1.**
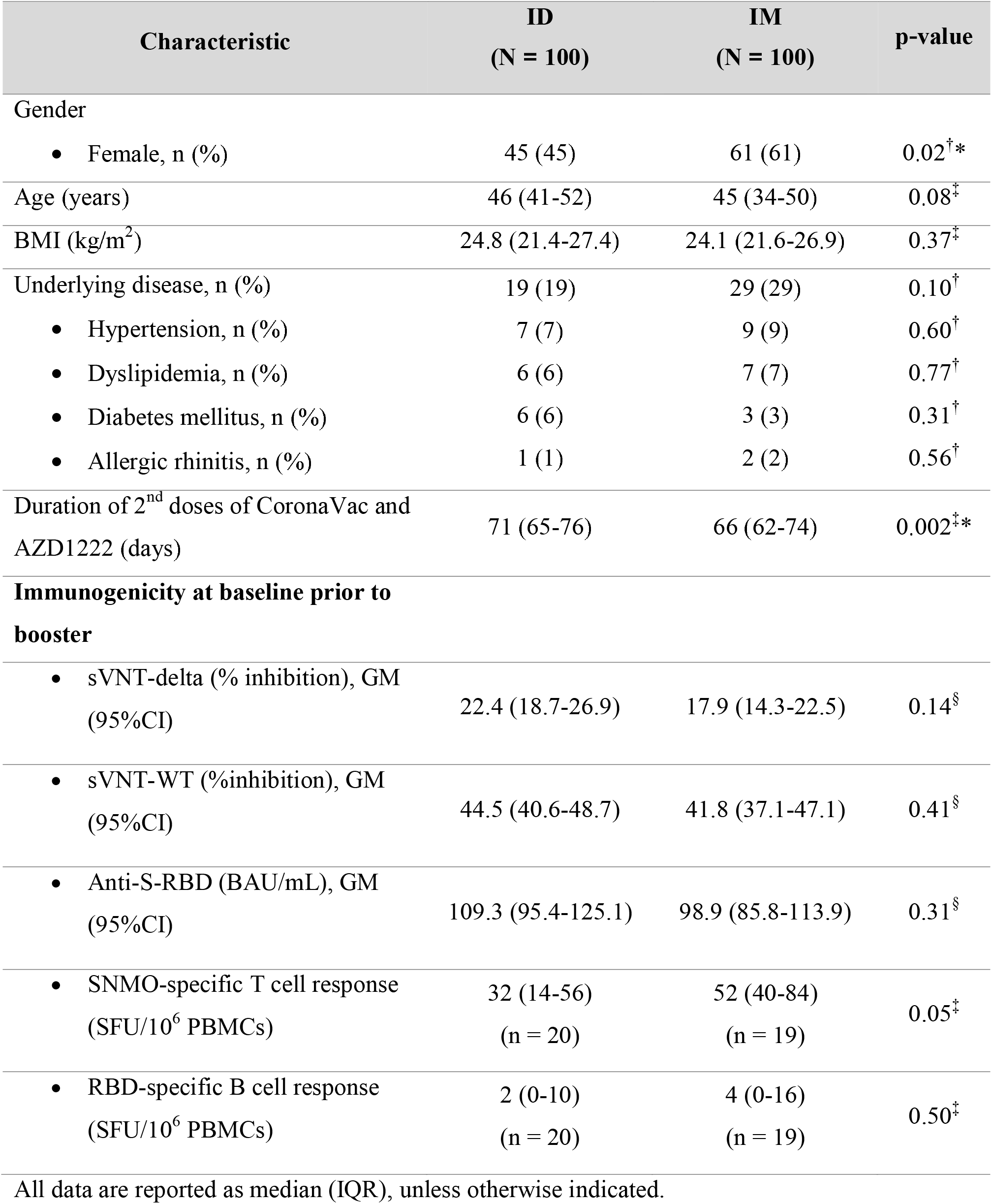
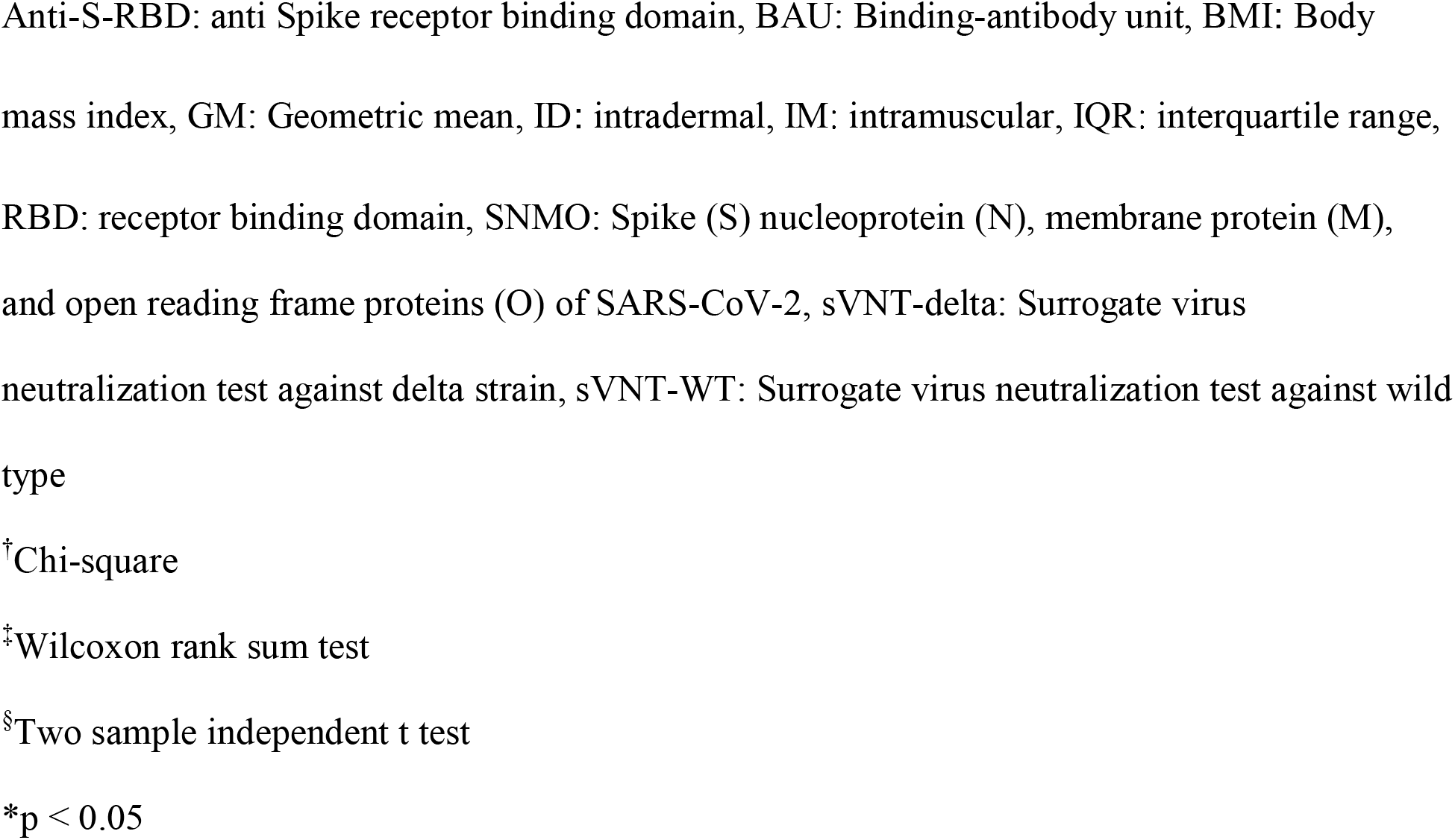
Baseline characteristics of participants receiving AZD1222 booster vaccine after 2-dose CoronaVac in healthy adults

| Characteristic | ID<br>(N = 100) | IM<br>(N = 100) | p-value |
| --- | --- | --- | --- |
| Gender |  |  |  |
| • Female, n (%) | 45 (45) | 61 (61) | 0.02<sup>†*</sup> |
| Age (years) | 46 (41-52) | 45 (34-50) | 0.08<sup>‡</sup> |
| BMI (kg/m<sup>2</sup>) | 24.8 (21.4-27.4) | 24.1 (21.6-26.9) | 0.37<sup>‡</sup> |
| Underlying disease, n (%) | 19 (19) | 29 (29) | 0.10<sup>†</sup> |
| • Hypertension, n (%) | 7 (7) | 9 (9) | 0.60<sup>†</sup> |
| • Dyslipidemia, n (%) | 6 (6) | 7 (7) | 0.77<sup>†</sup> |
| • Diabetes mellitus, n (%) | 6 (6) | 3 (3) | 0.31<sup>†</sup> |
| • Allergic rhinitis, n (%) | 1 (1) | 2 (2) | 0.56<sup>†</sup> |
| Duration of 2<sup>nd</sup> doses of CoronaVac and AZD1222 (days) | 71 (65-76) | 66 (62-74) | 0.002<sup>‡*</sup> |
| **Immunogenicity at baseline prior to booster** |  |  |  |
| • sVNT-delta (% inhibition), GM (95%CI) | 22.4 (18.7-26.9) | 17.9 (14.3-22.5) | 0.14<sup>§</sup> |
| • sVNT-WT (%inhibition), GM (95%CI) | 44.5 (40.6-48.7) | 41.8 (37.1-47.1) | 0.41<sup>§</sup> |
| • Anti-S-RBD (BAU/mL), GM (95%CI) | 109.3 (95.4-125.1) | 98.9 (85.8-113.9) | 0.31<sup>§</sup> |
| • SNMO-specific T cell response (SFU/10<sup>6</sup> PBMCs) | 32 (14-56)<br>(n = 20) | 52 (40-84)<br>(n = 19) | 0.05<sup>‡</sup> |
| • RBD-specific B cell response (SFU/10<sup>6</sup> PBMCs) | 2 (0-10)<br>(n = 20) | 4 (0-16)<br>(n = 19) | 0.50<sup>‡</sup> |

All data are reported as median (IQR), unless otherwise indicated.
Anti-S-RBD: anti Spike receptor binding domain, BAU: Binding-antibody unit, BMI: Body mass index, GM: Geometric mean, ID: intradermal, IM: intramuscular, IQR: interquartile range, RBD: receptor binding domain, SNMO: Spike (S) nucleoprotein (N), membrane protein (M), and open reading frame proteins (O) of SARS-CoV-2, sVNT-delta: Surrogate virus neutralization test against delta strain, sVNT-WT: Surrogate virus neutralization test against wild type <sup>†</sup>Chi-square <sup>‡</sup>Wilcoxon rank sum test <sup>§</sup>Two sample independent t test \*p < 0.05

### 3.2 Reactogenicity

Most common solicited reactogenicity reported was localized at injection site such as erythema and pain, as shown in Figure 1 and Supplementary Table 1. More than half (53%) participants reported erythema, lasting for median duration of 4 (IQR 3-6) days, which was mostly grade 1. Pain at injection site was reported in 43% with median duration of 2 (IQR 1-4) days and mostly grade 1. Other solicited reactogenicity reported were 40% fatigue, 30% myalgia, 27% headache, 18% feverish, 17% swelling, 12% arthralgia and 9% diarrhea. Vesicle and blister at injection site, which progressed to dry blister and turned to hyperpigmentation, were also reported as unsolicited reactogenicity (photo as shown in Supplementary Figure 1).

**Figure 1.**
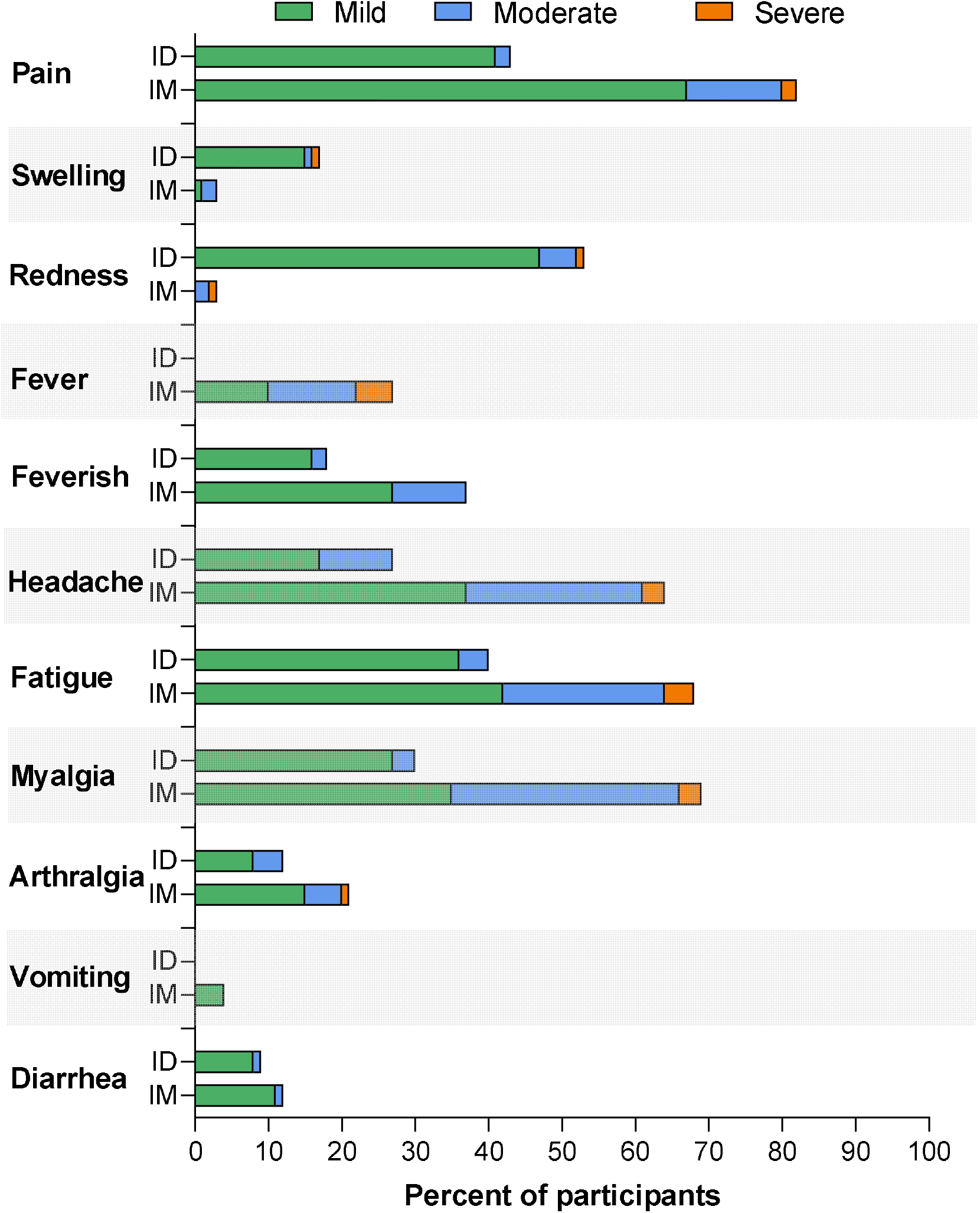
Solicited reactogenicity within 7 days of ID and IM AZD1222 booster after 2-dose CoronaVac in healthy adult. ID: intradermal, IM: intramuscular

Lower systemic reactogenicity including fever (0% ID versus 27% IM, p =N/A), feverish (18% ID versus 37% IM, p = 0.003), headache (27% ID versus 64% IM, p <0.001), fatigue (40% ID versus 68% IM, p <0.001), and myalgia (30% ID versus 69% IM, p <0.001) was reported in ID compared with IM, as shown in Supplementary Table 1.

### 3.3 Immunogenicity of ID booster

#### 3.3.1 sVNT

Ninety-eight percent of the participants achieved sVNT against delta strain and wild type ≥80%inhibition at both day 14 and day 28. The GMs (95% CI) of sVNT to delta strain were 95.5% inhibition (94.2-96.8) at day 14, and 93.7% inhibition (91.9-95.5) at day 28 after ID booster, as shown in Figure 2 and Table 2. The GMs (95% CI) of sVNT to wild type were 94.8% inhibition (94.0-95.6) at day 14, and 93.7% inhibition (92.1-95.4) at day 28, as shown in Table 2. At day 90 post booster vaccination, the GMs of sVNT, to both delta strain and wild type, waned to 73.1% inhibition (66.7-80.2) and 81.9% inhibition (76.2-88.0), respectively.

**Figure 2.**
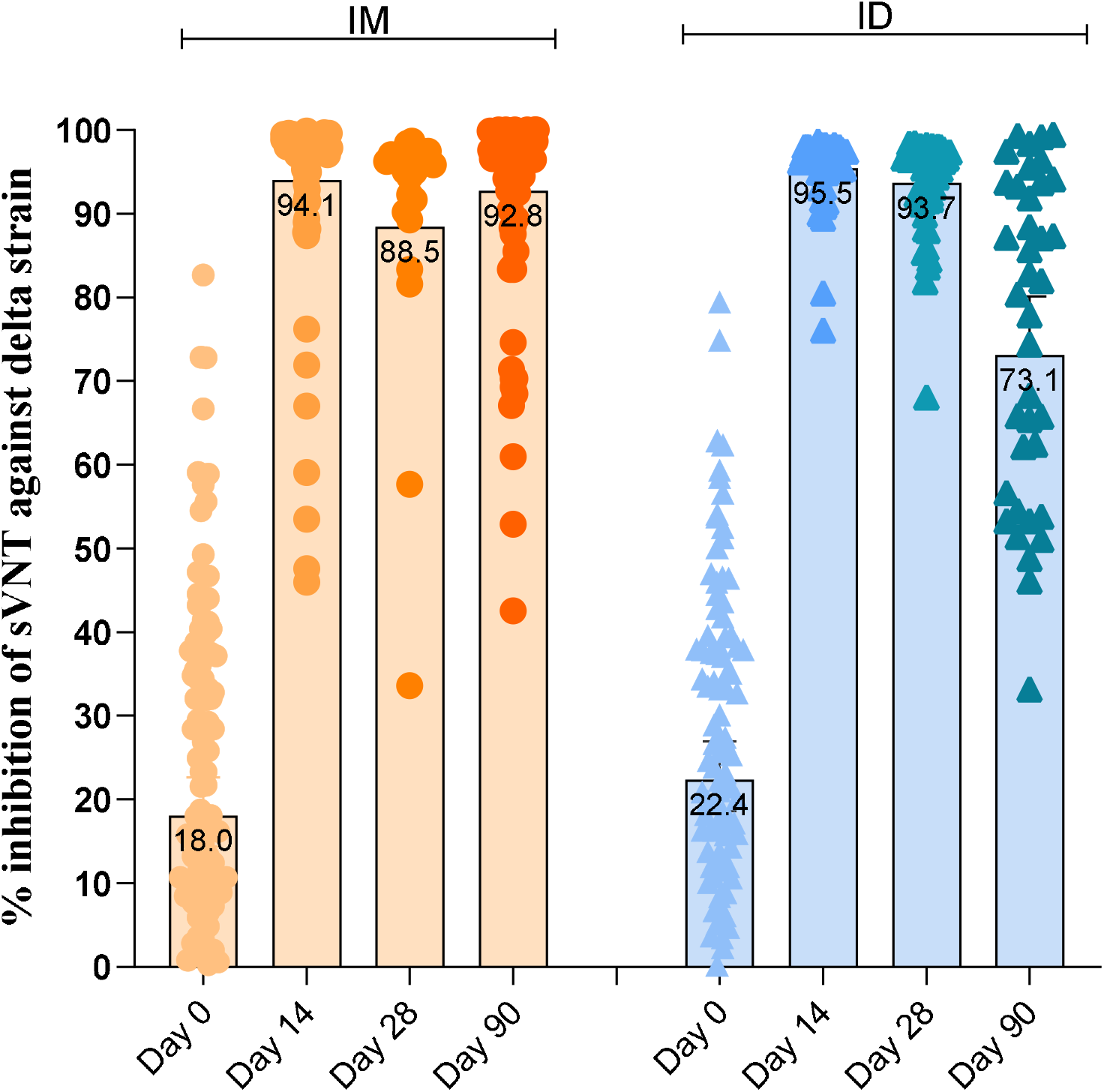
Comparison of sVNT % inhibition against delta strain at day 0, 14, 28, and 90 of ID and IM AZD1222 booster after 2-dose of CoronaVac in healthy adult. ID: intradermal, IM: intramuscular, sVNT: Surrogate virus neutralization test

**Table 2.**
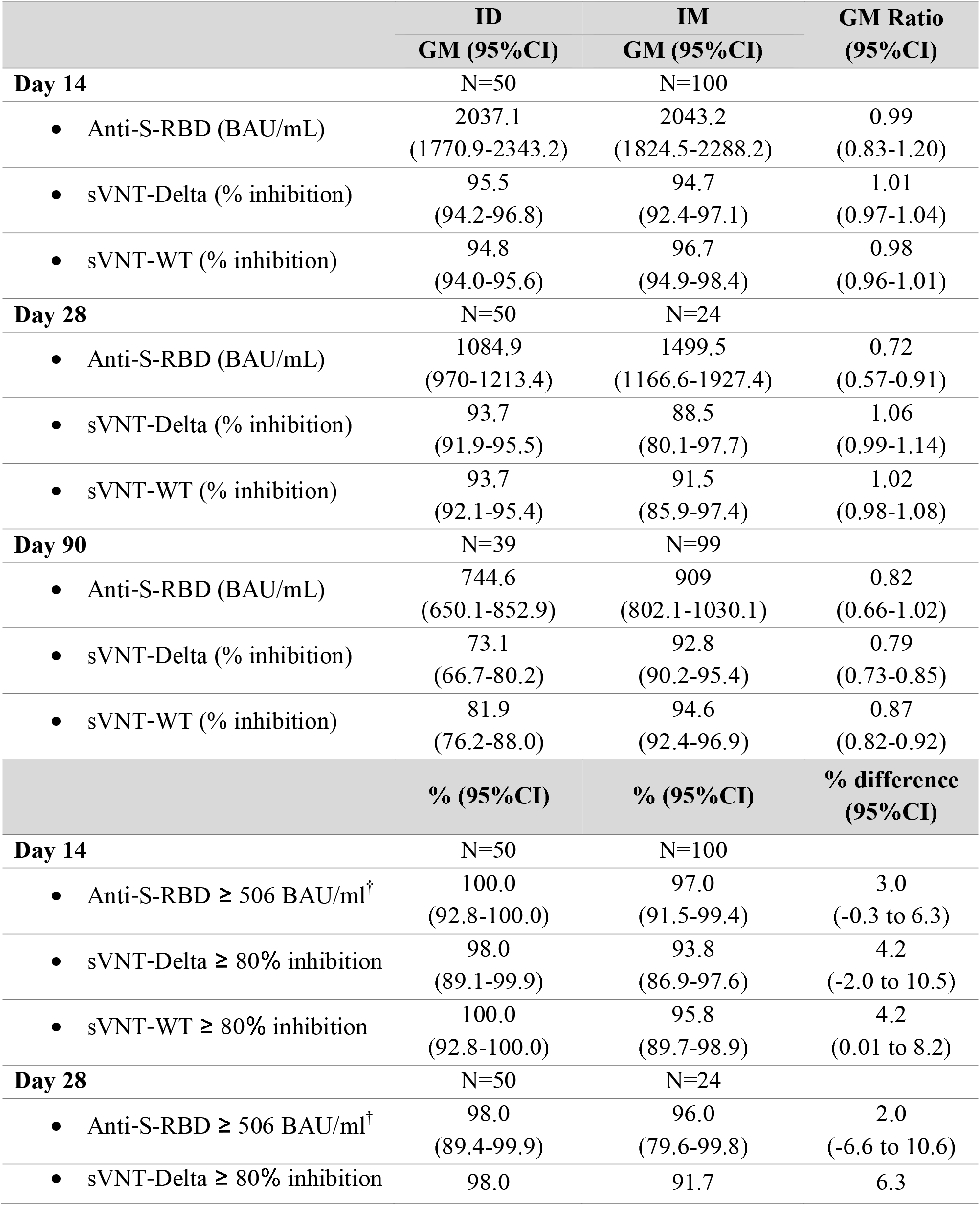
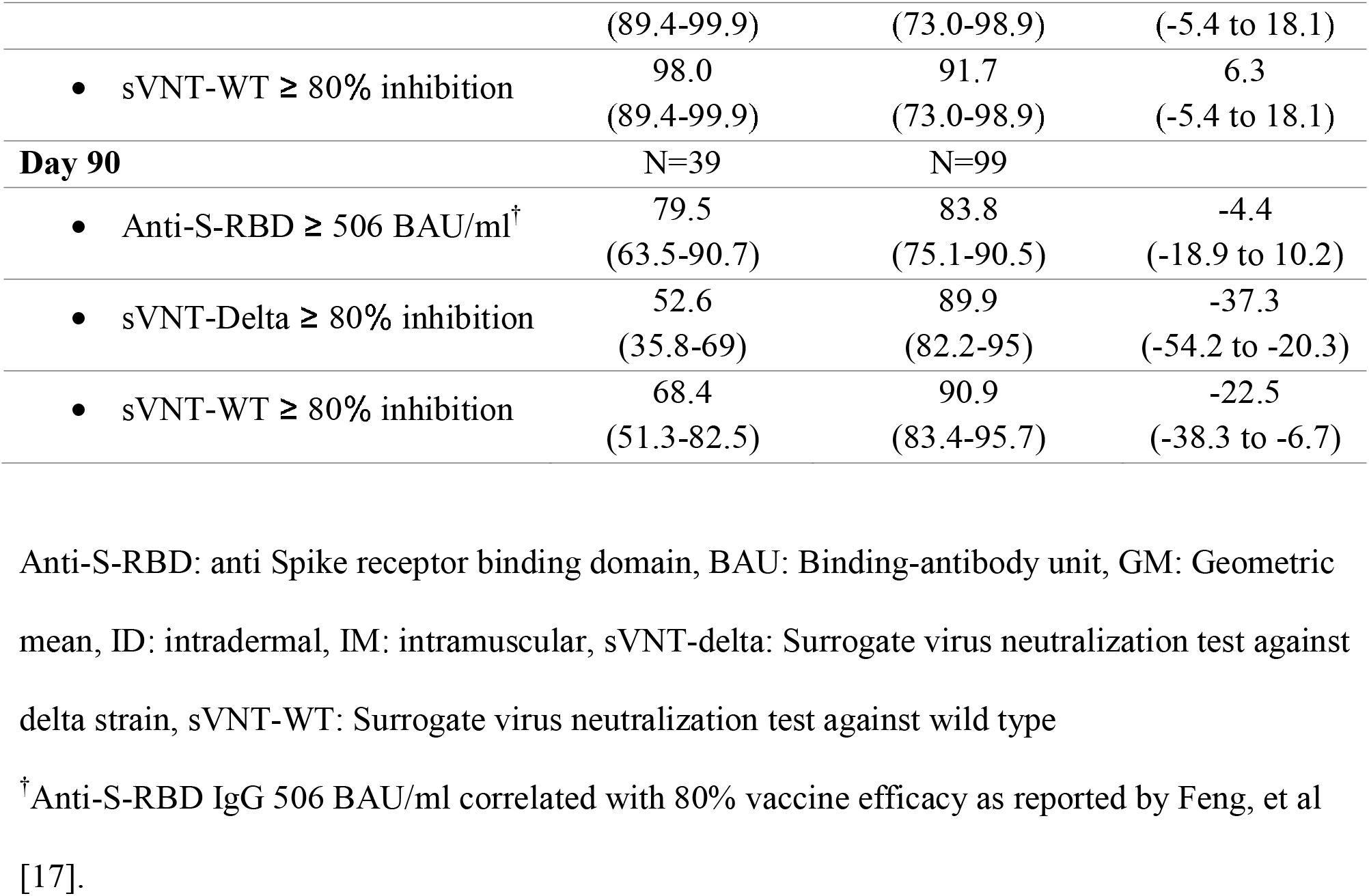
Comparison of intradermal and intramuscular AZD1222 booster immunogenicity in healthy adult completing 2-dose CoronaVac

#### 3.3.2 Anti-S-RBD IgG

The GMs (95% CI) of anti-S-RBD IgG were 2037.1 (1770.9-2343.2), 1084.9 (970.0-1213.4), and 744.6 (650.1-852.9) BAU/mL at day 14, 28, and 90 after ID booster, respectively, as shown in Table 2.

### 3.4 Immunogenicity of ID compared with IM booster

Proportion of participants with sVNT to delta strain ≥80% inhibition at day 14 was non-inferior among ID recipients compared with IM recipients, with difference of 4.2% (95% CI -2.0 to 10.5). But at day 90, it was significantly lower, with difference of -37.3% (−54.2 to −20.3). These differences were similar to sVNT to wild type, as shown in Table 2. GMR of anti-S-RBD IgG showed non-inferiority at day14, with GMR of 0.99 (0.83-1.20), and borderline inferior at day 90, with GMR of 0.82 (0.66-1.02). However, at 90 days after ID vaccination, only 79.5% had antibody exceeding the proposed protective level, compared with 83.8% of IM recipients.

### 3.5 T and B cell responses evaluated by ELISpot assay

From the sub study analysis of CMI response, ELISpot assay showed significant rise of T cell and B cell response at day 28 and declined at day 90, as shown in Figure 3. Median (IQR) of IFN-γ-producing T cell spots specific to SNMO protein-derived peptide pools at day 0 was 32 (14-56), at day 28 was 146 (70-192), and at day 90 was 90 (20-140) SFU/10^6^ PBMCs, respectively. Compared with IM study, the median (IQR) was 52 (40-48) at day 0, 96 (44-128) at day 28, and 44 (32-72) SFU/10^6^ PBMCs at day 90, respectively. Median (IQR) of RBD-specific memory B cell spots at day 0 was 2 (0-10), increased to 18 (14-36) at day 28, and declined to 6 (4-20) SFU/10^6^ PBMCs at day 90. Compared with IM study of 4 (0-16) at day 0, 26 (16-32) at day 28, and 8 (4-16) SFU/10^6^ PBMCs at day 90.

**Figure 3.**
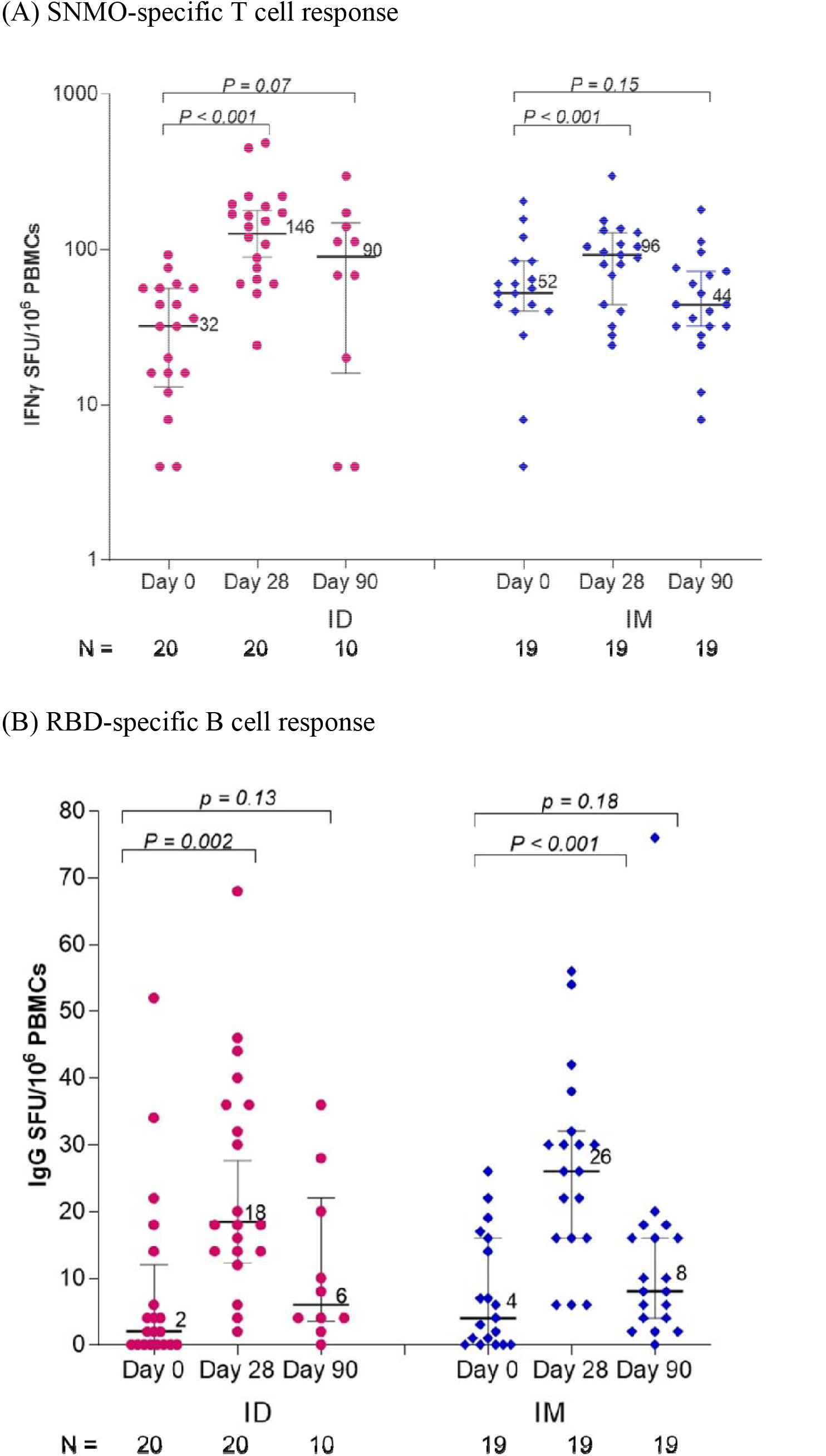
ELISpot assay at day 0, day 28, and day 90 of ID and IM AZD1222 booster in healthy adult completing 2-dose CoronaVac: (A) Interferon-γ ELISpot response to SNMO overlapping peptides of SARS-CoV-2, (B) RBD-specific IgG ELISpot assay. ID: intradermal, IM: intramuscular, RBD: receptor binding domain, SFU: spot forming unit, SNMO: Spike (S) nucleoprotein (N), membrane protein (M), and open reading frame proteins (O) of SARS-CoV-2

## 4. Discussion

ID AZD1222 booster vaccine in 2-dose-CoronaVac-primed adults raised high anti-S-RBD IgG >506 BAU/ml, and high levels of functional neutralizing antibodies >80% inhibition as measured by sVNT to wild type and delta strain, thus non-inferior to IM route at day 14. However, at 3 months post ID AZD1222 booster vaccination, this study demonstrated, despite similar anti-S-RBD IgG, but lower sVNT against delta strain to IM booster, suggesting more rapid waning neutralizing antibody response after ID compared to IM route. Most reactogenicity occurred locally with erythema, pain, and swelling at injection site. Erythema, swelling, and blister were reported more common in ID booster. Systemic symptoms such as fever, feverish, headache, fatigue, and myalgia were less common than conventional IM injection.

The non-inferior immunogenicity of ID vaccination was demonstrated for influenza, rabies, and hepatitis B vaccines [21]. Immunogenicity of ID AZD1222 at day 14 was not inferior to conventional IM booster vaccine as shown with GMR of anti-S-RBD and difference in proportion of participants having sVNT to delta strain and wild type passing 80%. This comparable result is similar to previous study in Netherland [22] which reported a robust antibody response from ID administration of mRNA COVID-19 vaccine at day 43 with comparable anti-spike IgG response for 10 µg ID with 100 µg IM mRNA-1273 vaccine. Additionally, recent report from Thailand also denoted a fractional-dose BNT162b2 ID booster, in healthy adults who had completed 2-dose inactivated vaccine for 2-3 months, induced comparable antibody level and function to the conventional IM booster when assessed on day 14 and 28 [14]. To our knowledge, no published report demonstrated immunogenicity results at 3 months post ID AZD1222 booster vaccination, which this study demonstrated inferior neutralizing antibodies.

Although, the importance of cellular immunity in correlation with vaccine protection is still unclear, specific T cells have been reported to reduce the severity of SARS-CoV-2 infection [23]. In this study, we have shown that a third dose of ID AZD1222 booster vaccine can increase specific T cell responses slightly higher than conventional IM route, similar to the previous report [24]. As opposed to previous ID BNT162b2 study, which failed to demonstrate T cell response, suggesting different vaccine platforms might play a role in cellular immune response. Specific memory B cells also have been reported to play a crucial role for effective responses to infection [25, 26]. Our result showed the slight boost of B cell response at 1 month and drop at 3 months, after ID AZD1222 booster. The timing of B cell study might be accounted for these responses, since the previous study showed the detectable B cell response after COVID-19 infection for 3-6 months [27].

This study reported local reactogenicity including erythema, blister, and pruritus after ID AZD1222 booster vaccine which is similar to previous report on rabies inactivated vaccine that more erythema and pruritus were reported from ID than conventional IM administration [28]. Also blister formation was reported after BCG vaccination that evolved over two weeks into an ulcer at injection site [29]. ID influenza vaccine study reported significant higher local adverse events particularly erythema and swelling, and also more common of fever and chills which is different from this study that fever was more common in IM vaccination [9]. Hyperpigmentation was also reported in this study as still seen on day 28 follow up visit which previous study of hepatitis B vaccine reported of local hyperpigmentation after ID vaccination in 55% [30]. Compared with parallel cohort IM study, ID booster had more local reactogenicity (erythema and swelling) at injection site but less pain and systemic reactogenicity (fatigue, myalgia, headache, feverish, arthralgia and diarrhea).

As current situation of COVID-19 pandemic, more vaccine supply is still needed for many countries as vaccine coverage is not enough to prevent mortality [31–33]. Almost half of the world population has received at least one dose of COVID-19 vaccine but only 2.2% of people in low-income countries have received vaccine [34]. AZD1222 or Astra Zeneca/Oxford COVID-19 vaccine has been used in Europe since December 2020 and also distributed in many countries including low to middle income countries [35]. As availability in many countries with limited vaccine supply, the ID administration of AZD1222 might be considered for mass vaccination as an advantage of dose-sparing technique [7]. However, there are some limitations, needs of skilled health providers for administration [36], and more rapid waning of neutralizing antibodies.

This study was limited by cohort study design without randomized control trial but there was the parallel cohort study with similar setting that should be able to benchmark the results. There were 2 factors that differed between the 2 groups. Specifically, there was more male in the ID group and the time interval between second and third dose was 1 week longer in the ID group. However, the immune responses at baseline before the third dose were comparable. Female was reported to have higher antibody response to vaccines [37] and after severe COVID- 19 [38]. The finding of later inferior neutralizing antibodies might be attributed to this gender difference, specifically more male participants in ID cohort. This study chose to determine the levels of functional neutralizing antibodies using the surrogate virus neutralization assay, rather than standard live-virus neutralization assay. However, we used the high cut-off value at 80% of sVNT in this study. Moreover, good correlations between sVNT and live-virus neutralization have been exhibited elsewhere [18, 39–41]. The strengths of this study were reporting complete solicited reactogenicity of all 100 participants with ID booster vaccination and multiple methods were used for immunity analysis including anti-S-RBD, sVNT (wild type and delta strain) and also CMI responses.

Intradermal AZD1222 booster vaccine in 2-dose-CoronaVac primed adult enhanced comparable short-term immunity, but inferior 3-month immunogenicity, with intramuscular administration. Reactogenicity was usually localized (erythema and pain) and less systemic than intramuscular vaccine. Due to more rapid waning neutralizing antibody, dose-sparing strategy with intradermal booster vaccination should be used in the setting of inadequate vaccine supply.

## Data Availability

All data produced in the present study are available upon reasonable request to the authors.

## Acknowledgements

Thank you for all study teams for their assistance with the study.

Chulalongkorn University Health Service Center

Santhiti Dahlan, M.D., Preeyanuch Panchim, Ratchadaporn Bumrungpipattanaporn, RN, Ampawan Sangchanpong, Wijitra Prayong

Department of Microbiology, Faculty of Medicine, Chulalongkorn University Asst.Prof.Pokrath Hansasuta, M.D., Vichaya Ruenjaiman, PhD, Supapit Horpratum, Jullada Thawilwang

Center of Excellence in Pediatric Infectious Diseases and Vaccines

Pintip Suchartlikitwong, M.D., Wipaporn Natalie Songtaweesin, M.D., Pathariya Promsena, M.D., Monta Tawan, RN, Jitthiwa Athipunjapong, RN, Thutsanun Meepuksom, RN, Angsumalin Sutjarit, RN, Juthamanee Moonwong, Rachaneekorn Nadsasarn, Thidarat Jupimai, Pornpavee Nuncharoen, Sasiprapha Khamthi, Pathomchai Amornrattanapaijit, Phattharapa Khamkhen,

National Center for Genetic Engineering and Biotechnology (BIOTEC)

Anan Jongkaewwattana, PhD, Kirana Yoohat, Channarong Seepiban, Jaraspim Narkpuk, Thorntun Deangphare,

Clinical Research Laboratory, Faculty of Medicine, Chulalongkorn University Palida Pingthaisong, Suwat Wongmueng

Faculty of Medicine, Thammasat University

Asst.Prof.Sira Nantapisal, M.D., Ph.D.

## Abbreviations

BAU: Binding-antibody unit
BMI: Body mass index
CMI: Cell-mediated immunity
ELISpot: Enzyme-linked immunospot
GM: Geometric mean
GMR: Geometric mean ratio
ID: Intradermal
IM: Intramuscular
PBMC: Peripheral blood mononuclear cell
SFU: Spot forming unit
S-RBD: Spike receptor binding domain
sVNT: Surrogate virus neutralization test

---

**Supplementary Figure 1.**
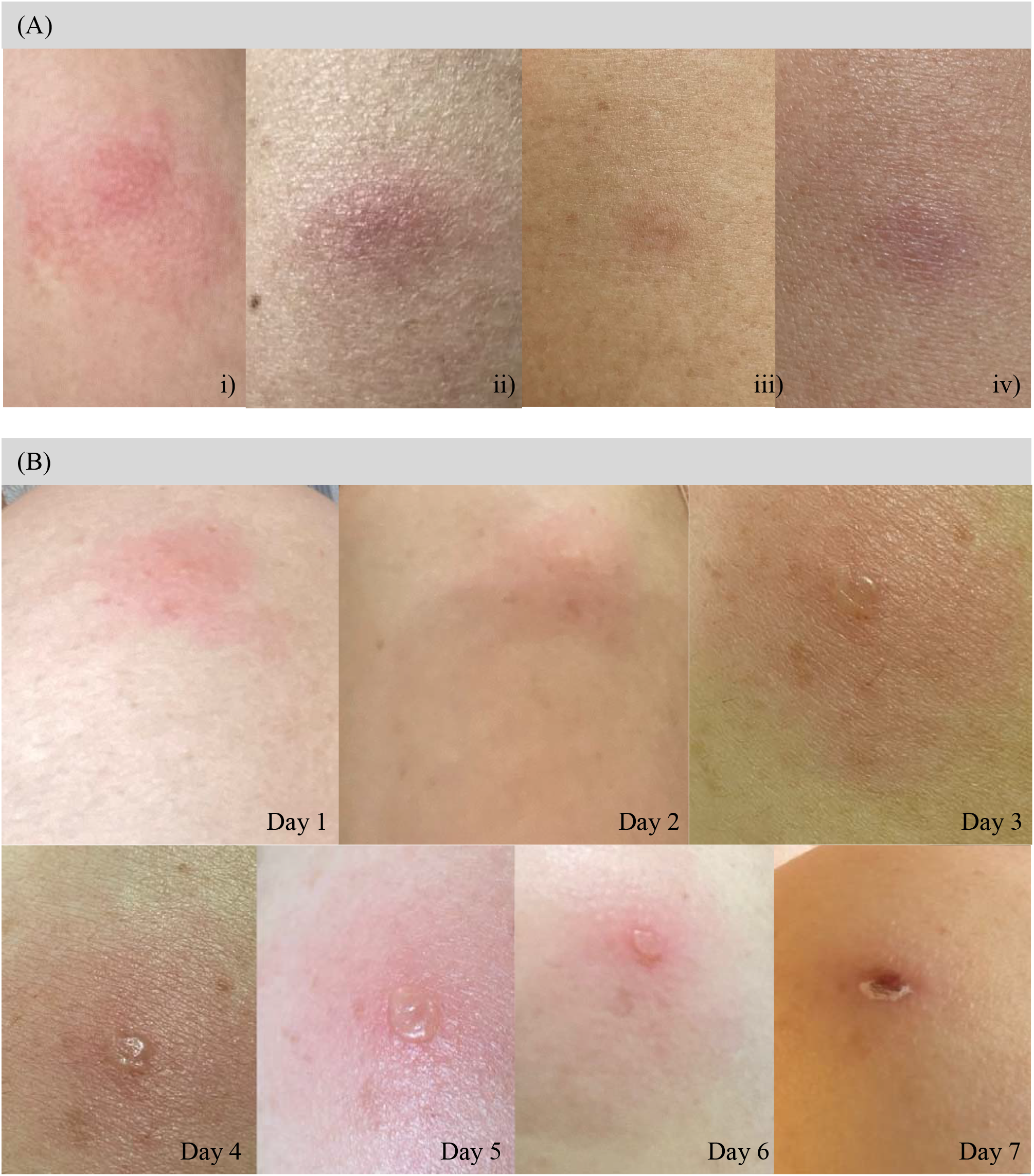
Photography of local reactogenicity in participants receiving intradermal AZD1222 booster after completing 2-dose CoronaVac: (A) local reactogenicity in participants: i-ii) erythematous patch at injection site at day 4, iii) residual post inflammatory hyperpigmented macule and iv) erythematous patch at injection site at day 28 and (B) a serial photography day 1-7 in single participant: Day 1 erythematous patch at injection site, Day 2 central swelling on erythematous patch, Day 3-6 blister forming at central of erythematous patch, and Day 7 dry blister with necrotic crust on erythematous base.

**Supplementary Table 1.**
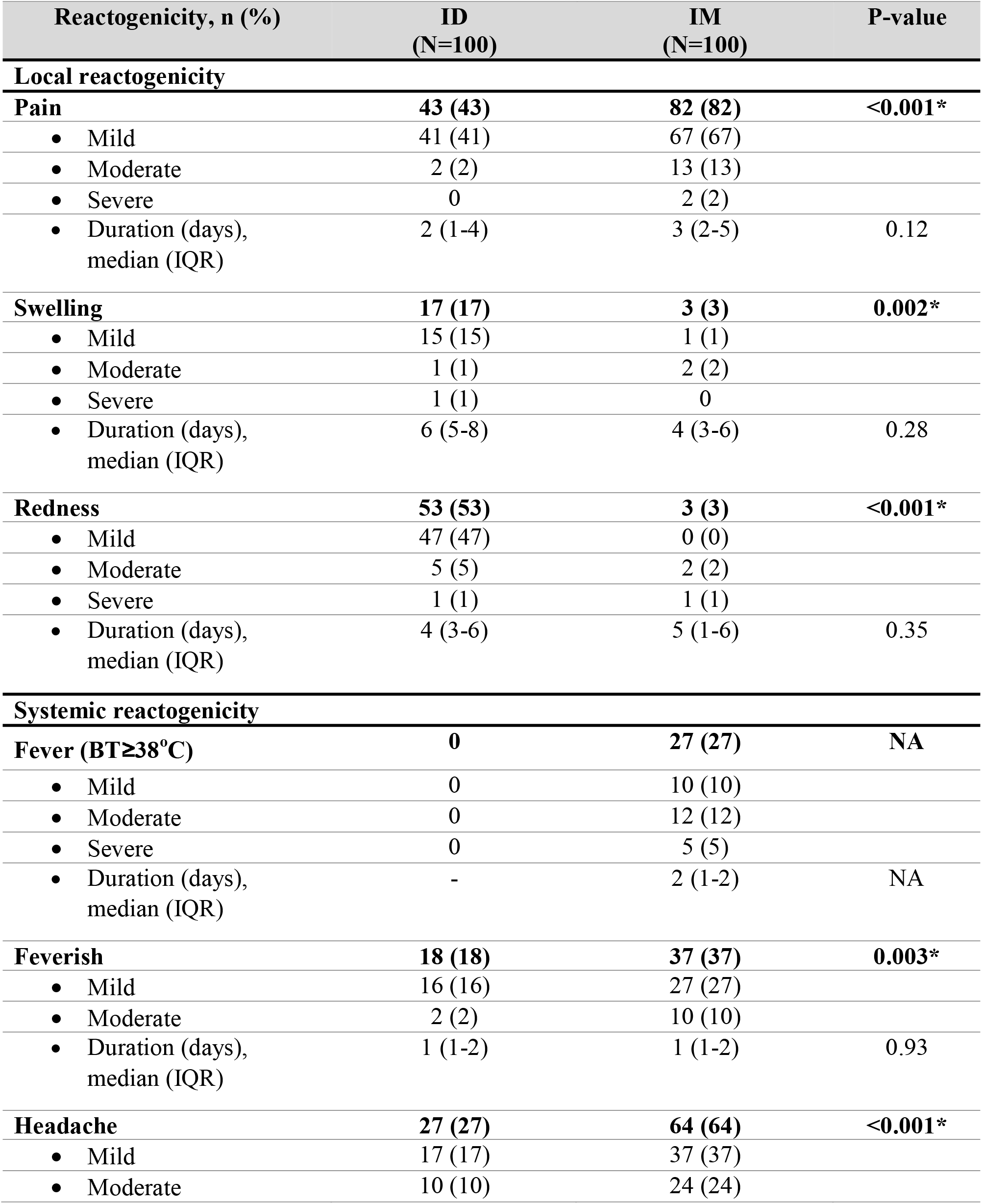
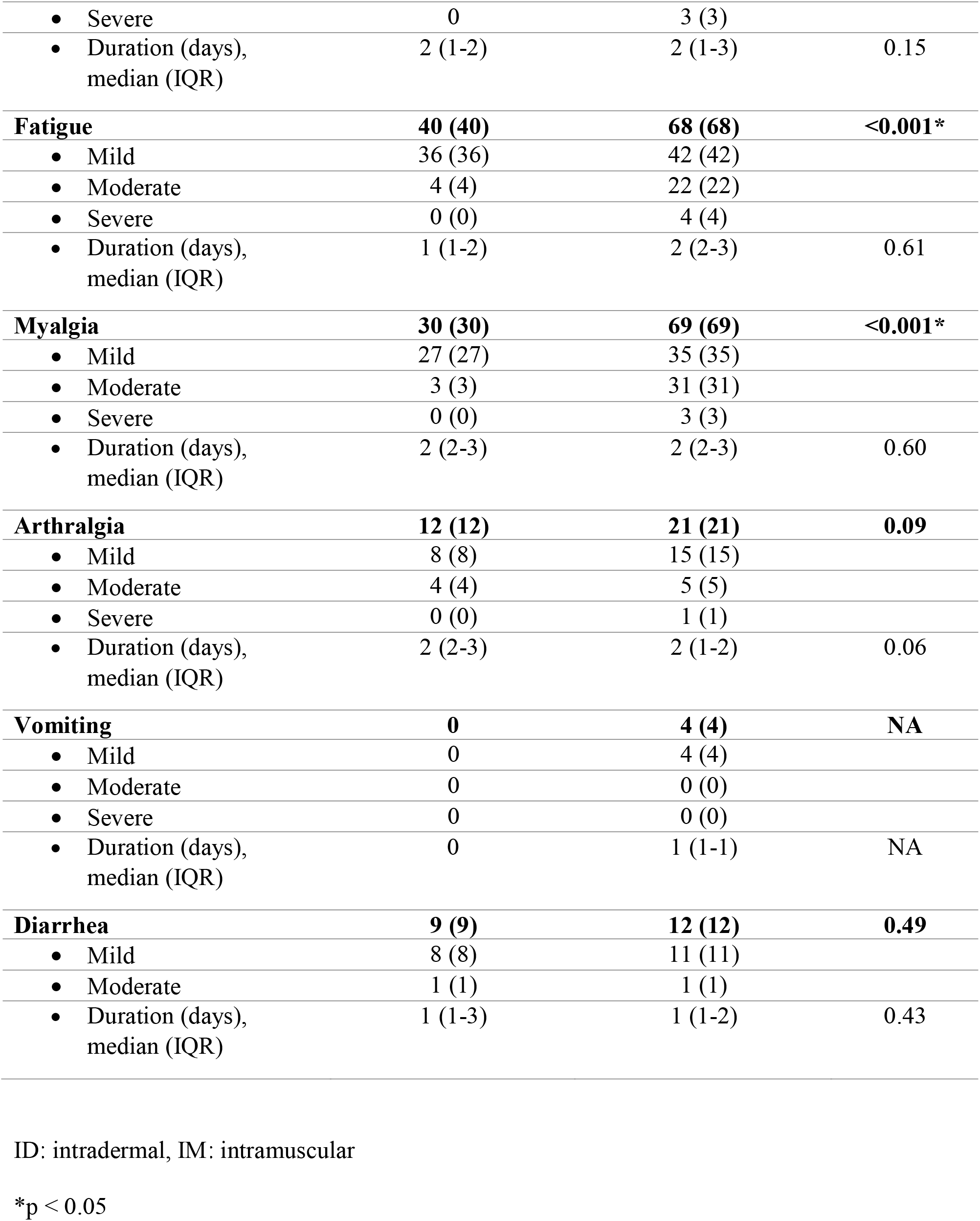
Solicited reactogenicity within 7 days of ID and IM AZD1222 booster after 2-dose CoronaVac in healthy adult.

| Reactogenicity, n (%) | ID<br>(N=100) | IM<br>(N=100) | P-value |
| --- | --- | --- | --- |
| **Local reactogenicity** |  |  |  |
| **Pain** | **43 (43)** | **82 (82)** | **<0.001\*** |
| • Mild | 41 (41) | 67 (67) |  |
| • Moderate | 2 (2) | 13 (13) |  |
| • Severe | 0 | 2 (2) |  |
| • Duration (days),<br>median (IQR) | 2 (1-4) | 3 (2-5) | 0.12 |
| **Swelling** | **17 (17)** | **3 (3)** | **0.002\*** |
| • Mild | 15 (15) | 1 (1) |  |
| • Moderate | 1 (1) | 2 (2) |  |
| • Severe | 1 (1) | 0 |  |
| • Duration (days),<br>median (IQR) | 6 (5-8) | 4 (3-6) | 0.28 |
| **Redness** | **53 (53)** | **3 (3)** | **<0.001\*** |
| • Mild | 47 (47) | 0 (0) |  |
| • Moderate | 5 (5) | 2 (2) |  |
| • Severe | 1 (1) | 1 (1) |  |
| • Duration (days),<br>median (IQR) | 4 (3-6) | 5 (1-6) | 0.35 |
| **Systemic reactogenicity** |  |  |  |
| **Fever (BT≥38°C)** | **0** | **27 (27)** | **NA** |
| • Mild | 0 | 10 (10) |  |
| • Moderate | 0 | 12 (12) |  |
| • Severe | 0 | 5 (5) |  |
| • Duration (days),<br>median (IQR) | - | 2 (1-2) | NA |
| **Feverish** | **18 (18)** | **37 (37)** | **0.003\*** |
| • Mild | 16 (16) | 27 (27) |  |
| • Moderate | 2 (2) | 10 (10) |  |
| • Duration (days),<br>median (IQR) | 1 (1-2) | 1 (1-2) | 0.93 |
| **Headache** | **27 (27)** | **64 (64)** | **<0.001\*** |
| • Mild | 17 (17) | 37 (37) |  |
| • Moderate | 10 (10) | 24 (24) |  |
| • Severe | 0 | 3 (3) |  |
| • Duration (days),<br>median (IQR) | 2 (1-2) | 2 (1-3) | 0.15 |
| **Fatigue** | **40 (40)** | **68 (68)** | **<0.001\*** |
| • Mild | 36 (36) | 42 (42) |  |
| • Moderate | 4 (4) | 22 (22) |  |
| • Severe | 0 (0) | 4 (4) |  |
| • Duration (days),<br>median (IQR) | 1 (1-2) | 2 (2-3) | 0.61 |
| **Myalgia** | **30 (30)** | **69 (69)** | **<0.001\*** |
| • Mild | 27 (27) | 35 (35) |  |
| • Moderate | 3 (3) | 31 (31) |  |
| • Severe | 0 (0) | 3 (3) |  |
| • Duration (days),<br>median (IQR) | 2 (2-3) | 2 (2-3) | 0.60 |
| **Arthralgia** | **12 (12)** | **21 (21)** | **0.09** |
| • Mild | 8 (8) | 15 (15) |  |
| • Moderate | 4 (4) | 5 (5) |  |
| • Severe | 0 (0) | 1 (1) |  |
| • Duration (days),<br>median (IQR) | 2 (2-3) | 2 (1-2) | 0.06 |
| **Vomiting** | **0** | **4 (4)** | **NA** |
| • Mild | 0 | 4 (4) |  |
| • Moderate | 0 | 0 (0) |  |
| • Severe | 0 | 0 (0) |  |
| • Duration (days),<br>median (IQR) | 0 | 1 (1-1) | NA |
| **Diarrhea** | **9 (9)** | **12 (12)** | **0.49** |
| • Mild | 8 (8) | 11 (11) |  |
| • Moderate | 1 (1) | 1 (1) |  |
| • Duration (days),<br>median (IQR) | 1 (1-3) | 1 (1-2) | 0.43 |

ID: intradermal, IM: intramuscular
\*p < 0.05

